# Influence of the hospital upon the outcomes of gastrointestinal fistulas: results of a multi-continent, multi-national, multi-center cohort

**DOI:** 10.1101/2021.10.20.21265268

**Authors:** Humberto Arenas Márquez, María Isabel Turcios Correia, Juan Francisco García, Roberto Anaya Prado, Arturo Vergara, Jorge Luis Garnica, Alejandra Cacho, Daniel Guerra, Miguel Mendoza Navarrete, Sergio Santana Porbén

## Abstract

**Rationale:** Prognosis and outcomes of gastrointestinal fistulas (GIF) might depend upon the operational characteristics of the hospital containing and caring for the patients.

**Objective:** To assess how selected operational characteristics of the hospital participating in the exercises of the “Fistula Day” Project (FDP) influence upon prognosis and outcomes of GIF.

**Study design:** Cohort-type study. Enrolled patients were followed for 60 days. Three cross-sectional examinations were made during the completion of the exercises of the FDP, namely, upon admission of the patient in the study, and 30 and 60 days after admission.

**Study serie:** Seventy-six hospitals of Latin America (13 countries) and Europe (4).

**Methods:** Associations between survival of the patient, prolongation of hospital stay, and (likely) spontaneous closure of the fistula, on one hand; and selected operational characteristics of the participating hospital, on the other; were assessed.

**Results:** Specialties hospitals prevailed. Most of the hospitals assisted between 1 – 2 GIF patients a month. Participating hospitals distributed evenly regarding the number of beds. Most of the hospitals had an intensive care unit. Similarly, three-quarters of the hospitals had a multidisciplinary unit dedicated to clinical and hospital nutrition. However, a unit dedicated to the management of intestinal failure and/or postoperative fistulas was present only in a fifth of them. Experience of the physician attending GIF was rated between “Expert” and “High” in one third of the hospitals. Number of hospitals beds associated with increased survival of GIF patients (χ^2^ = 5.997; p = 0.092), prolonged hospital stay (χ^2^ = 7.885; p < 0.05), and higher rate of spontaneous closure of the fistula (χ^2^ = 11.947; p < 0.05). In addition, rate of spontaneous closure of the fistula was (marginally) higher among patients assisted by a hospital unit specialized on intestinal failure (χ^2^ = 3.610; p = 0.0574). On the other hand, survival of the patient was dependent (also marginally) upon the number of patients assisted in a month (χ^2^ = 5.934; p = 0.0514).

**Conclusions:** It is likely number of hospital beds to determine prognosis and outcomes of GIF. Other operational characteristics of the hospital might exert a marginal influence upon survival of the patient and the likely spontaneous closure of the fistula.

## INTRODUCTION

The “Fistula Day” Project has as supraobjective the improvement of the care provided to patients complicated with a gastrointestinal fistula (GIF) in Latin American (LATAM) hospitals. To that end, the aforementioned project foresees the conduction of surveys on a regular basis that might serve to collect current evidences on the current care of GIF, evolution and progression of the patients, and the nutritional and surgical practices adopted as part of the containment and resolution of GIF.

Results observed with a first cohort of 177 patients surveyed in 76 LATAM and European hospitals were presented in a preceding essay.^1^ Type of fistula influenced upon prognosis and evolution of the GIF patient: patients with an enteroatmospheric fistula (EAF) showed a lower survival (and complementarily a higher mortality). On the other hand, hospital stay was prolonged in patients subjected to elective surgery as well as those with a reduced calf circumference. Location of fistula also influenced upon prolongation of hospital stay.

The results annotated with the studied cohort have prompted the “Fistula Day” investigators to consider other determinants of the prognosis and outcomes of GIF. In that sense, it is likely operational characteristics of the hospital to influence upon resolution of GIF, being the treatment and management of GIF a complex clinical-surgical situation and thus demanding specialized surgical, pharmaceutical and nutritional care. If this is to be the case, recommendations might be pertinent regarding the choice of the hospital better endowed for treating and containing GIF, the equipment and engineering of the hospitals for dealing successfully with GIF, and training the medical care teams in all the procedures they require. Given all the aforementioned, this work has been conducted in order to examine the influence of selected operational characteristics of the hospital participating in the “Fistula Day” Project upon prognosis and outcomes of GIF.

## MATERIAL AND METHOD

The design of the “Fistula Day” Project has been previously presented.^1^ In brief, hospitals eventually included in the project were invited to submit demographic, clinical, sanitary, surgical and nutritional data of patients complicated with a GIF between the months of May 2018 and July 2018 (both included) in three consecutive surveys.^1^ Hospitals also submitted data regarding their operational characteristics such as: type of hospital (General *vs*. Specialties), number of patients cared | treated for a fistula in a month work (1 – 2 patients, 3 – 4 patients, 5 and more patients); and number of beds: Between 1 – 100 beds, 101 – 200 beds, 201 – 300 beds, 301 – 400 beds, 401 – 500 beds; and > 500 beds.

Participant hospitals were also surveyed regarding the inclusion within the organigram of the institution of an Intensive Care Unit (ICU), a multidisciplinary unit dedicated to Clinical nutrition, and a unit dedicated to the treatment of intestinal failure and/or postoperative fistula (IFU). In addition, hospitals were asked about the experience of the acting physician regarding the treatment of intestinal leakages/fistulas: Expert Grade, High, Medium, Low, and None.

### Data processing and statistical-mathematical analysis of results

Data submitted by the hospitals involved in the “Fistula Day” project were entered into an on line application built with RedCap®©^*^ (University of Vanderbilt, United States). R program for statistical management and analysis (R Core Team 2018 version 3.5.0, United States) was used for debugging, preparing and processing the data collected during the “Fistula Day” surveys. Data were reduced down to absolute | relative frequencies and percentages according with the type of variable and the purpose of the statistical analysis.

Condition of the patient upon discharge (Alive/Deceased), prolongation of hospital stay (Yes/No) and spontaneous closure of the GIF (Yes/No) were assumed as the 30- and 60-days outcomes of the “Fistula Day” project. Nature and strength of the associations between the “Fistula Day” outcomes on one hand, and the operational characteristics of the hospitals included in the study cohort, on the other, were assessed by means of appropriate statistical tests in accordance with the type of the variable. Differences existing between cohorts of patients given the selected predictor were assessed by means of the *log-rank* test based on the chi-square distribution.^2^ A level < 5 % was used in all instances to denote the finding as significant.

### Treatment of missing data

Data missed during follow-up of the patient were replaced with the observation annotated in the preceding cross-sectional examination in accordance with the “Last Observation Carried Forward” (LOCF) principle.

### Intention-to-treat

Data gathered during the “Fistula Day” project was analyzed according with the “Intention to treat” principle in order to keep the size of the cohort constant.^3^

### Ethical considerations

The protocol followed by local surveyors during the “Fistula Day” was drafted according with “Good Clinical Practices”.^3^ Identity and rights of the surveyed patients were also protected.^4^ The patients (and by extension their caregivers) were informed about the purposes of the research, and the non-invasive nature of the procedures. Collected data were adequately preserved in order to ensure anonymity and confidentiality. Aggregated data were used in the interpretation of the results and the realization of statistical inferences. Informed consent was obtained from the patient before inclusion in the cohort. Local conduction of the activities foreseen in the “Fistula Day” was authorized and supervised by the hospital Committees of Ethics after presentation, review and approval of the research protocols.

The researchers entrusted with the conduction of the “Fistula Day Project” presented the protocol “Current status of the postoperative fistula of digestive tract; multicentric, multinational study. DAY OF THE FISTULA” before the Ethics Committee of the San Javier Hospital (city of Guadalajara, State of Jalisco, México) for review and approval. A ruling was emitted on April 11^th^, 2018 by Dr. Eduardo Razón Gutiérrez, acting Director of the Ethics Committee, with the approval of the research protocol and the authorization for the conduction of the “Fistula Day” Project^†^.

## RESULTS

At the conclusion of the “Fistula Day” Project the indicators of GIF evolution and prognosis were as follows:^1^ *Survival*: 85.3 %; *Prolonged hospitalization*: 46.3 %; and *Spontaneous closure of fistula*: 36.2 %.

### Characteristics of the participating hospitals

Table 1 shows the operational characteristics of the participating hospitals. Seventy-six hospitals were involved in the “Fistula Day” activities. Specialties hospitals prevailed (at least numerically). Most of the hospitals assisted between 1 – 2 GIF patients in a month work. Participating hospitals distributed evenly regarding number of beds. Most of the hospitals had an ICU. Similarly, three-quarters of the hospitals had a multidisciplinary unit dedicated to Clinical and hospital nutrition. On the contrary, a unit dedicated to the treatment of intestinal failure and/or postoperative fistulas was present in only a quarter of the hospitals. Experience of the acting physician in GIF management was rated between “Expert” and “High” in a third of the hospitals.

**Table 1.** Operational characteristics of the hospitals participating in the “Fistula Day” Project. Number and [within brackets] percentages of hospitals observed in the corresponding category of the characteristic are shown.

| Characteristic | Findings |
| --- | --- |
| Type of hospital | General: 32 [42.1]<br>Specialties: 44 [57.9] |
| Number of patients assisted/treated for fistulas in a month work | 1 – 2: 43 [56.6]<br>3 – 4: 12 [15.8]<br>≥ 5: 21 [27.6] |
| Number of beds in the participating hospitals | 1 – 100: 13 [17.1]<br>101 – 200: 16 [21.1]<br>201 – 300: 15 [19.7]<br>301 – 400: 8 [10.5]<br>401 – 500: 8 [10.5]<br>> 500: 16 [21.1] |
| Intensive Care Unit included within the participating hospital | Present: 72 [94.7]<br>Absent: 4 [ 5.3] |
| Multidisciplinary unit dedicated to Clinical nutrition within the participating hospital | Present: 59 [77.6]<br>Absent: 17 [22.4] |
| Unit dedicated to the management of intestinal failure and/or postoperative fistula included within the participating hospital | Present: 22 [28.9]<br>Absent: 54 [71.1] |
| Experience of the attending physician in the management of intestinal leakages/fistulas | Expert: 12 [15.8]<br>High: 36 [23.7]<br>Medium: 18 [23.7]<br>Low: 6 [ 7.9]<br>None: 4 [ 5.3] |

Source: Records of the study.
Size of the study serie: 76.

### Influence of the hospital upon survival of the patient

Table 2 shows the associations between the operational characteristics of the participating hospital and survival of the patient. Of all the examined characteristics, only the number of GIF patients assisted in a month work influenced (albeit marginally) upon survival of the patient (χ^2^ = 5.934; p = 0.0514; test of independence based on the chi-square distribution). Association between the number of patients assisted in the hospital, on one hand, and survival, on the other; adopted an inverted “U” shape: *1 – 2 patients a month*: 85.9 % (Δ = +0.7 % regarding global, non-adjusted, survival rate); *3 – 4 patients a month*: 97.1 % (Δ = +11.8 %); and ≥ *5 patients a month*: 79.2 % (Δ = -6.1 %).

**Table 2.** Associations between survival of the patient at the study closure and the characteristics of the participating hospitals. Number and [within brackets] percentages of patients observed in the corresponding category of the characteristic are shown. Legend: ICU: Intensive Care Unit. MCNU: Multidisciplinary Clinical Nutrition Unit. IFU: Intestinal Failure Unit.

| Characteristic | Findings | Interpretation |
| --- | --- | --- |
| *Survival of the patient* |  |  |
| Type of hospital | General: 86.7<br>Specialties: 84.7 | $\chi^2 = 0.090$ |
| Patients with fistulas assisted in the hospital | 1 – 2/months: 85.9<br>3 – 4/months: 97.1<br>≥ 5/months: 79.2 | $\chi^2 = 5.934$<br>p = 0.0514 |
| Number of hospital beds | 1 – 100 beds: 72.7<br>101 – 200 beds: 93.9<br>201 – 300 beds: 84.4<br>301 – 400 beds: 93.7<br>401 – 500 beds: 81.0<br>> 500 beds: 84.9 | $\chi^2 = 5.997$ |
| Hospital ICU | Present: 85.3<br>Absent: 85.7 | $\chi^2 = 0.0009$ |
| MCNU | Present: 83.8<br>Absent: 91.4 | $\chi^2 = 1.303$ |
| IFU | Present: 83.3<br>Absent: 86.7 | $\chi^2 = 0.378$ |
| Experience of the attending physician | Expert: 82.7<br>High: 83.7<br>Medium: 85.3<br>Low: 89.5<br>None/Non declared: 85.7 | $\chi^2 = 0.464$ |

Source: Records of the study. Size of the study serie: 177.

Figure 1 shows the behavior of the survival of the patient in each of the moments of the study cohort. None of the characteristics of the hospital influenced upon the survival rate observed in each moment.

**Figure 1.**
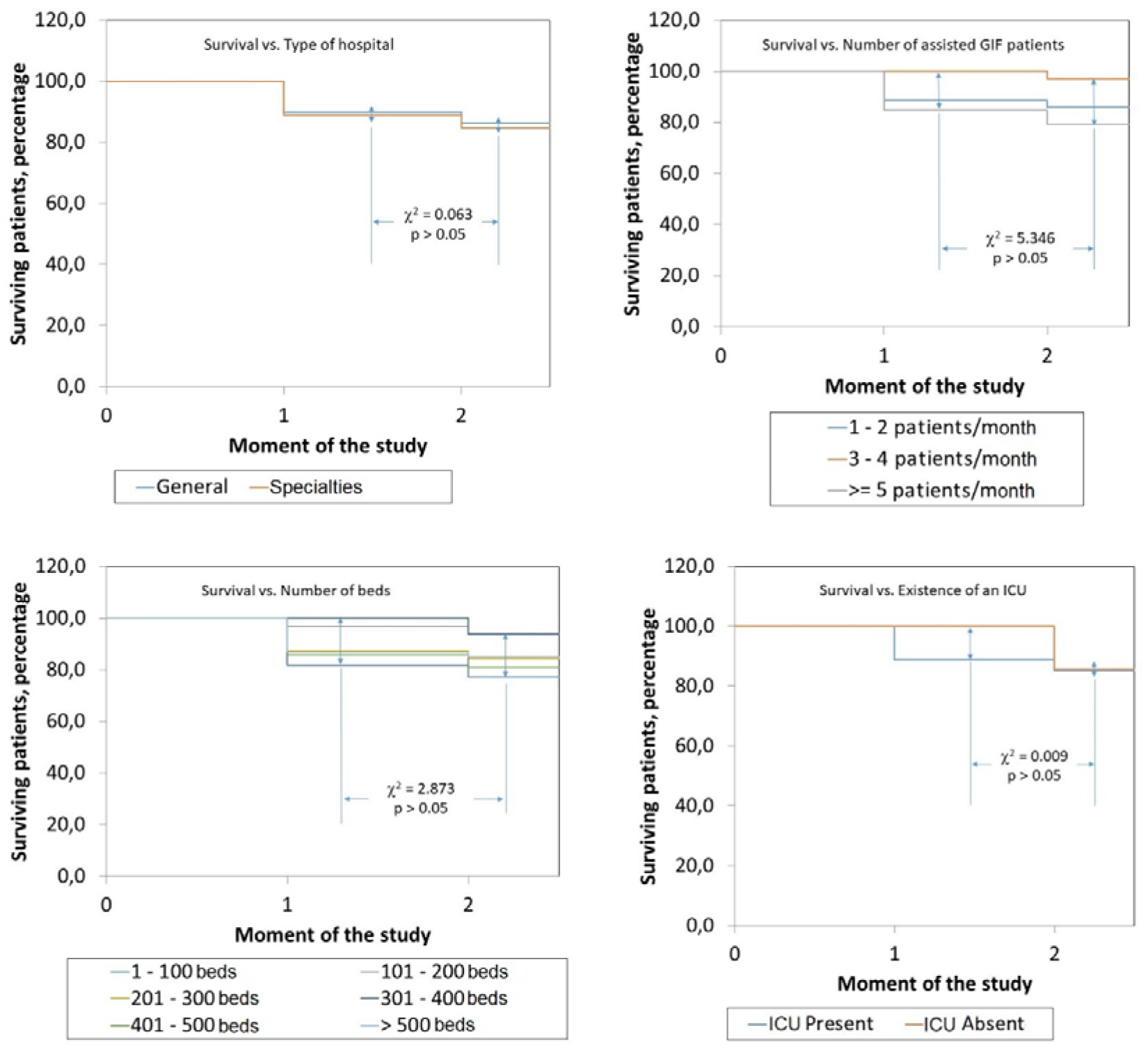
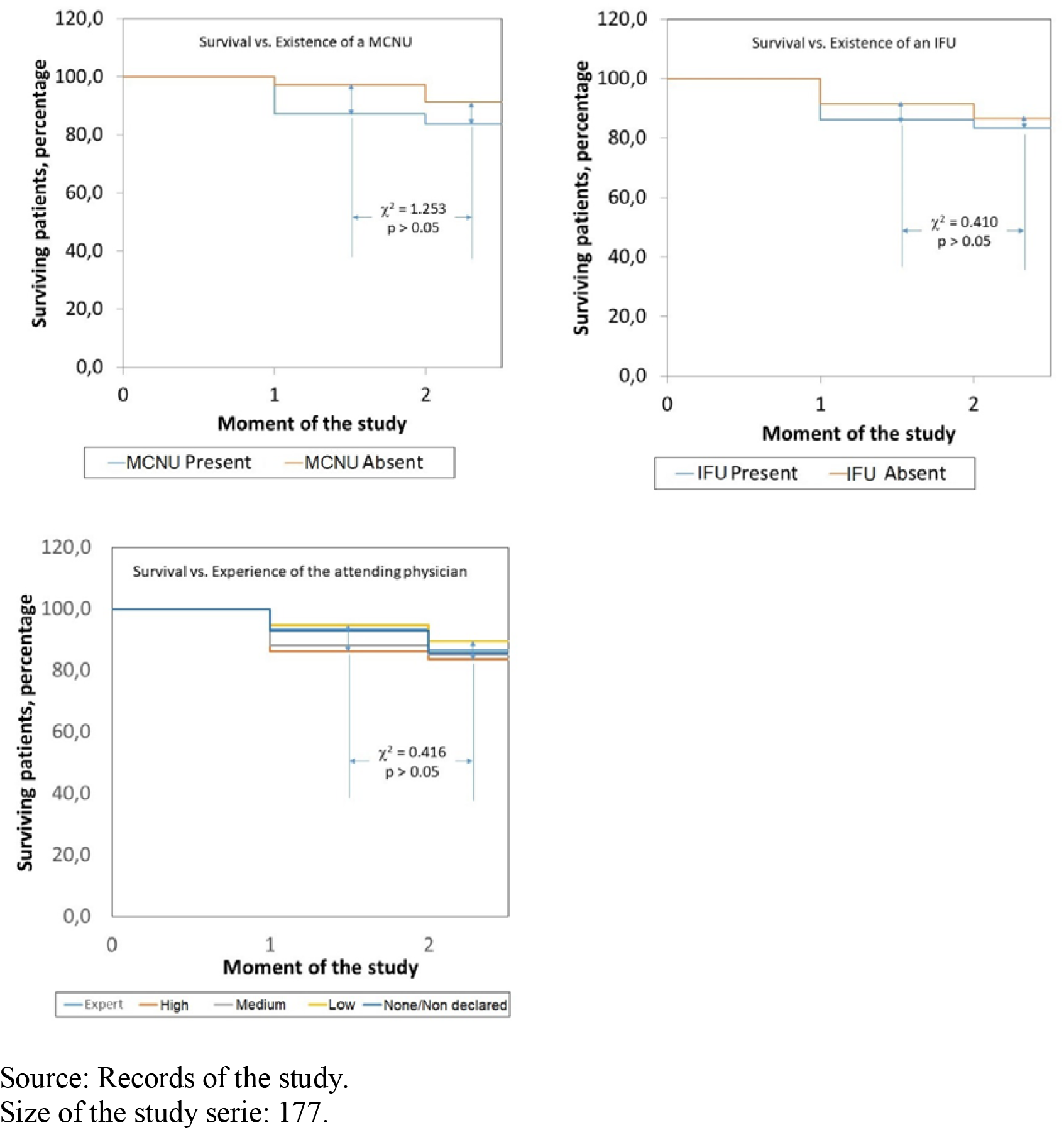
Influence of the operational characteristics of the participating hospitals upon the survival of patients with gastrointestinal fistulas in each moment of the study. Legend: GIF: Gastrointestinal fistulas. ICU: Intensive Care Unit. MCNU: Multidisciplinary Clinical Nutrition Unit. IFU: Intestinal Failure Unit.

### Influence of the hospital upon prolongation of hospitalization of the patient

Table 3 shows the associations between the characteristics of the participating hospital and prolonged hospitalization of the patient. Among all the examined characteristics, only the number of hospital beds stood out as a predictor of the prolonged hospitalization of the patient (χ^2^ = 19.267; test of independence based on the chi-square distribution). Association between the number of hospital beds, on one hand, and prolonged hospitalization, on the other; adopted a “U” shape: the highest rate of prolonged hospitalizations was observed in hospitals with as many as 100 beds as well as those with > 500 beds: ≤ *100 beds*: 72.7 % of the patients (Δ = +26.4 % regarding the global, non-adjusted rate) *vs. > 500 beds*: 43.4 % (Δ = -2.9 %; p < 0.05).

**Table 3.**
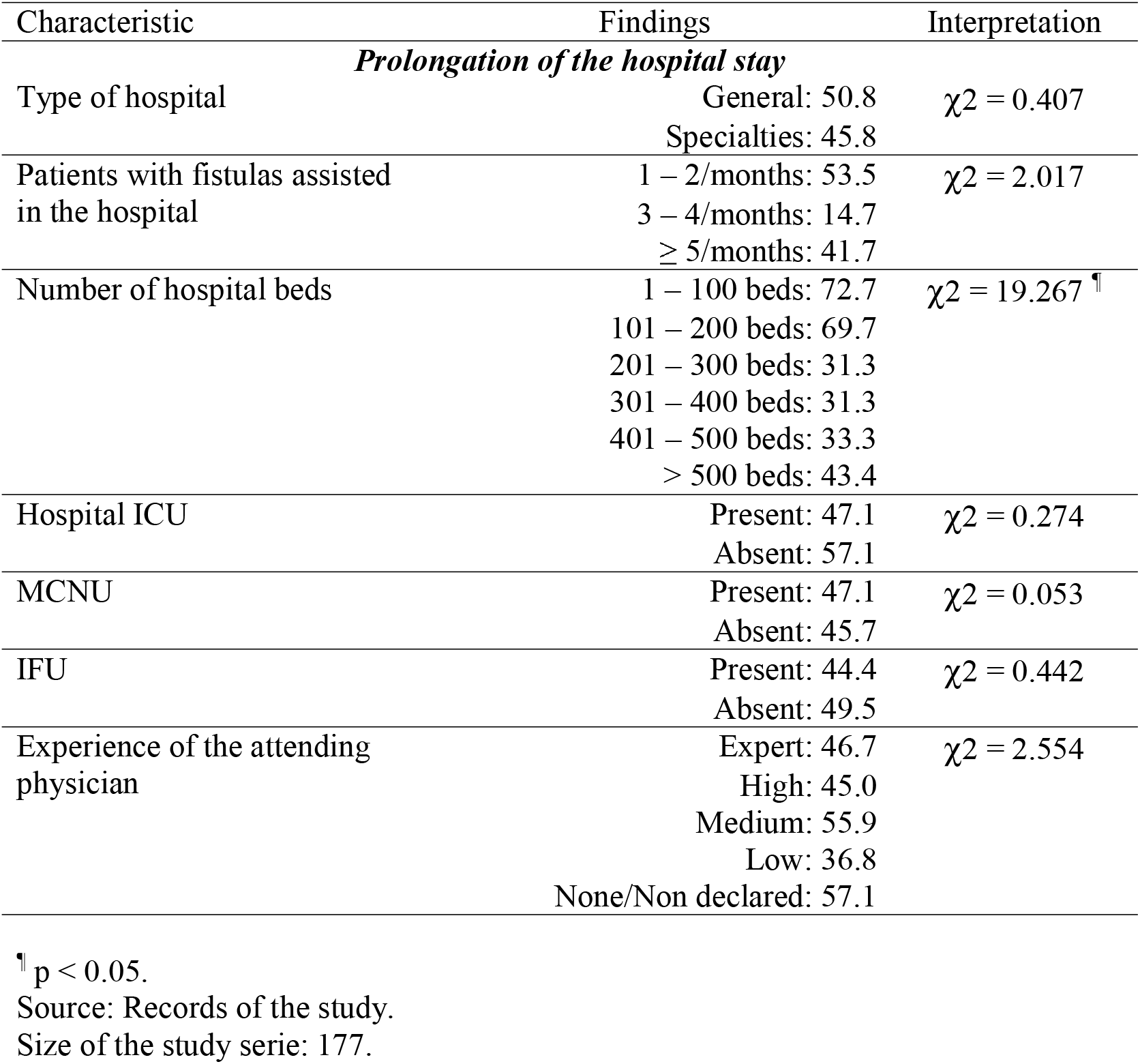
Associations between prolongation of the hospital stay at the study closure and the characteristics of the participating hospitals. Number and [within brackets] percentages of patients observed in the corresponding category of the characteristic are shown. Legend: ICU: Intensive Care Unit. MCNU: Multidisciplinary Clinical Nutrition Unit. IFU: Intestinal Failure Unit.

| Characteristic | Findings | Interpretation |
| --- | --- | --- |
| ***Prolongation of the hospital stay*** |  |  |
| Type of hospital | General: 50.8<br>Specialties: 45.8 | $\chi^2 = 0.407$ |
| Patients with fistulas assisted in the hospital | 1 – 2/months: 53.5<br>3 – 4/months: 14.7<br>≥ 5/months: 41.7 | $\chi^2 = 2.017$ |
| Number of hospital beds | 1 – 100 beds: 72.7<br>101 – 200 beds: 69.7<br>201 – 300 beds: 31.3<br>301 – 400 beds: 31.3<br>401 – 500 beds: 33.3<br>> 500 beds: 43.4 | $\chi^2 = 19.267^{\text{¶}}$ |
| Hospital ICU | Present: 47.1<br>Absent: 57.1 | $\chi^2 = 0.274$ |
| MCNU | Present: 47.1<br>Absent: 45.7 | $\chi^2 = 0.053$ |
| IFU | Present: 44.4<br>Absent: 49.5 | $\chi^2 = 0.442$ |
| Experience of the attending physician | Expert: 46.7<br>High: 45.0<br>Medium: 55.9<br>Low: 36.8<br>None/Non declared: 57.1 | $\chi^2 = 2.554$ |

<sup>¶</sup>p < 0.05.
Source: Records of the study.
Size of the study serie: 177.

Figure 2 shows the behavior of prolonged hospitalization of the patient in each of the moments of the cohort. Similar to the aforementioned, only the number of hospital beds influenced upon the behavior of the cohort of patients: in each moment of the cohort, the hospitals with the smallest number of beds distinguished themselves for the highest rates of prolonged hospitalizations (χ^2^ = 6.180; p < 0.05; log-rank test based on the chi-square distribution).

**Figure 2.**
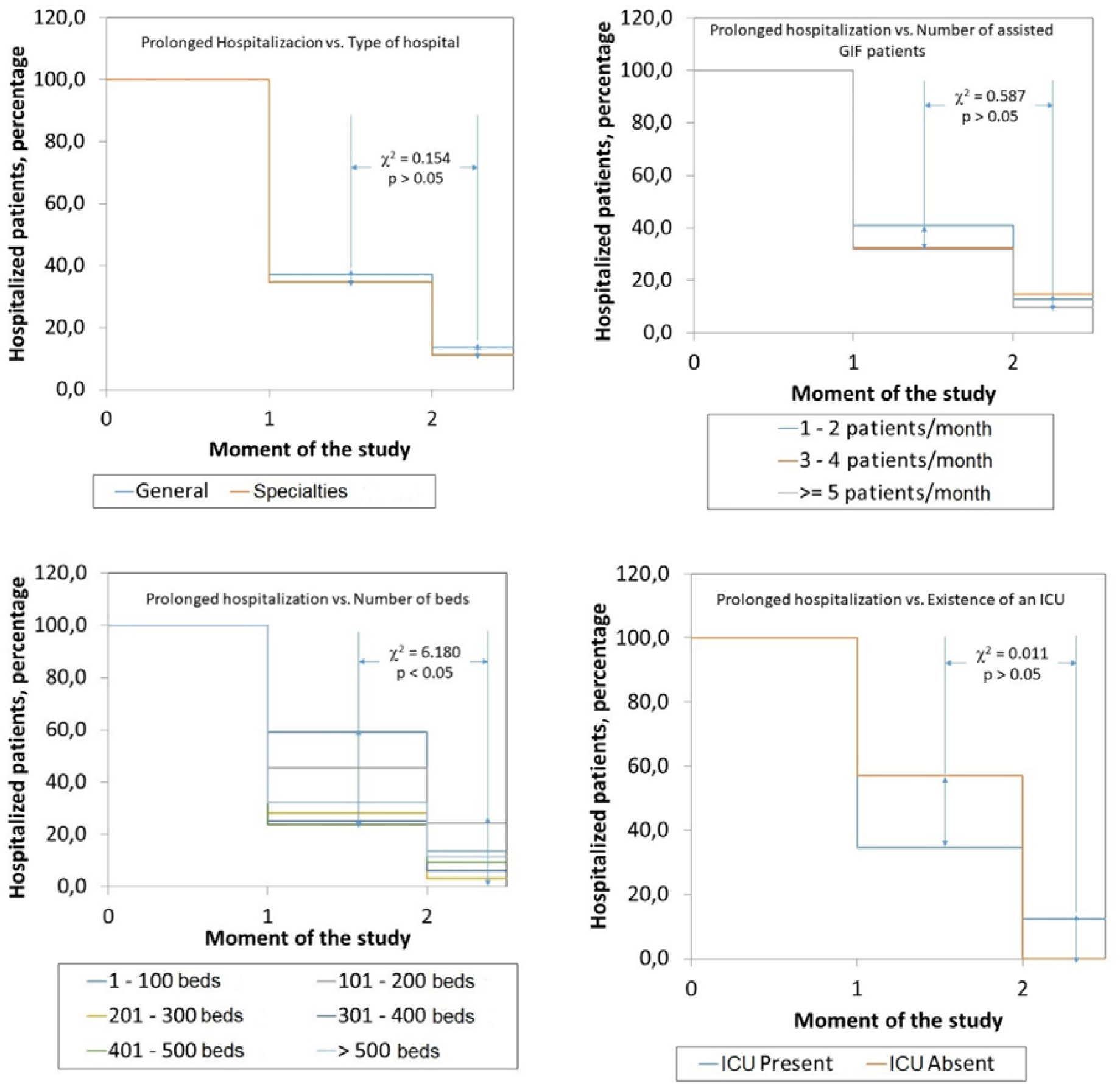
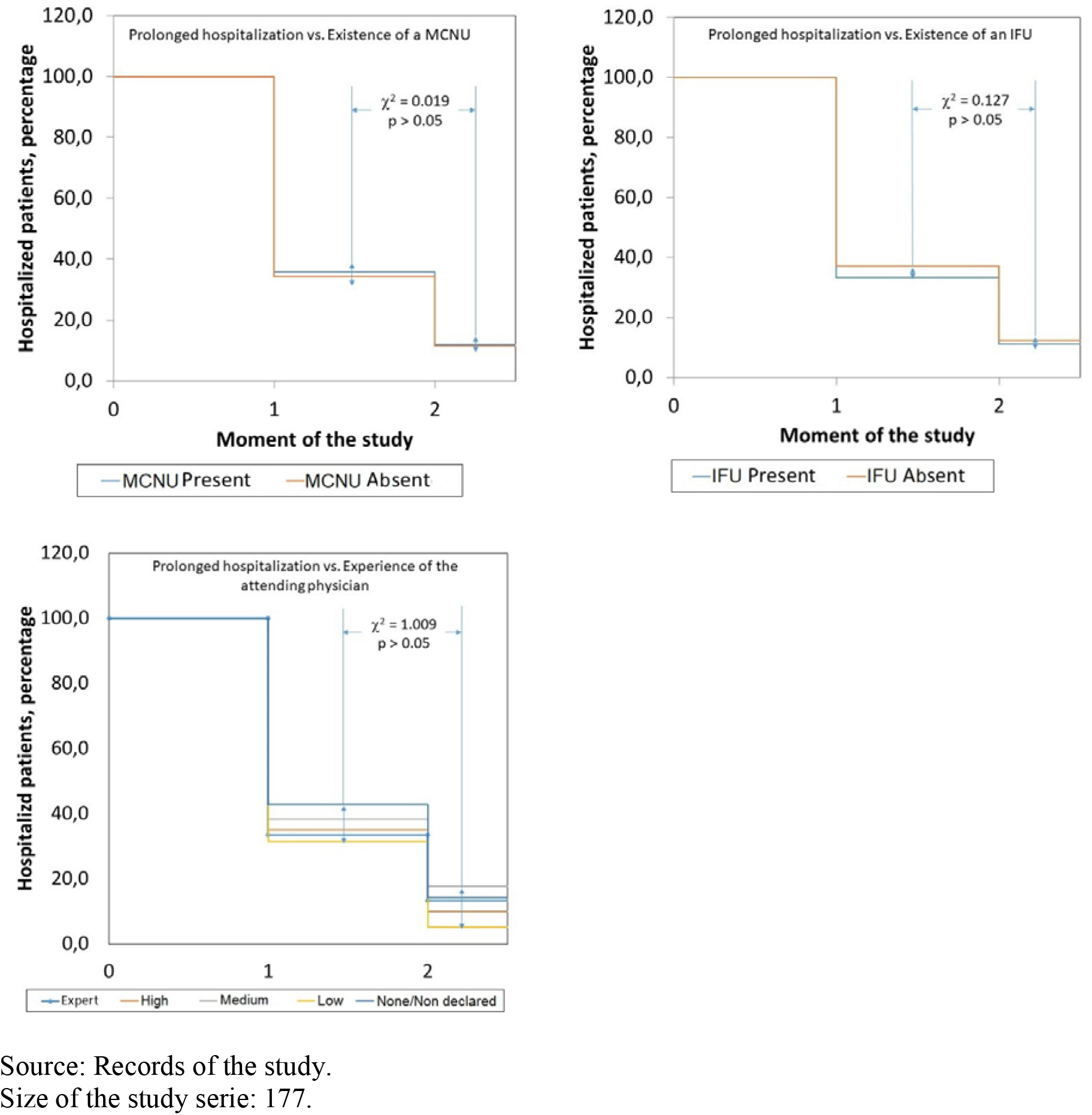
Influence of the operational characteristics of the participating hospitals upon the prolongation of hospital stay in each moment of the study. Legend: GIF: Gastrointestinal fistulas. ICU: Intensive Care Unit. MCNU: Multidisciplinary Clinical Nutrition Unit. IFU: Intestinal Failure Unit.

### Influence of the hospital upon the spontaneous closure of the fistula

Finally, Table 4 shows the influence of the hospital upon the likelihood of spontaneous closure of the GIF. Of all the examined characteristics, number of beds was also the only one influencing upon the spontaneous closure of the fistula (χ^2^ = 11.947; test of independence based on the chi-square distribution). Hospitals with the highest number of beds showed the highest rate of spontaneous closure of the fistula: ≤ *100 beds*: 13.6 % (Δ = -22.6 % with respect to global, non-adjusted rate); *Between 101 – 200 beds*: 51.5 % (Δ = +15.3 %); *Between 201 – 300 beds*: 28.1 % (Δ = -8.1 %); *Between 301 – 400 beds*: 25.0 % (Δ = -11.2 %); *Between 401 – 500 beds*: 33.3 % (Δ = -2.9 %); and *> 500 beds*: 45.3 % (Δ = +9.1 %); respectively (χ^2^ = 11.947; p < 0.05; test of independence based on the chi-square distribution). It is to be noticed existence of an IFU influenced (albeit marginally) upon the possibility of the spontaneous closure of GIF: existence of an IFU in the hospital pointed to a higher number of spontaneous closures (p = 0.0574; test of independence based upon the chi-square distribution).

**Table 4.** Associations between spontaneous closure of the gastrointestinal fistulas at the study closure and the characteristics of the participating hospitals. Number and [within brackets] percentages of patients observed in the corresponding category of the characteristic are shown. Legend: ICU: Intensive Care Unit. MCNU: Multidisciplinary Clinical Nutrition Unit. IFU: Intestinal Failure Unit.

| Characteristic | Findings | Interpretation |
| --- | --- | --- |
| *Spontaneous closure of the fistula* |  |  |
| Type of hospital | General: 33.9<br>Specialties: 37.3 | $\chi^2 = 0.195$ |
| Patients with fistulas assisted in the hospital | 1 – 2/month: 31.0<br>3 – 4/months: 35.3<br>≥ 5/months: 41.7 | $\chi^2 = 1.780$ |
| Number of hospital beds | 1 – 100 beds: 13.6<br>101 – 200 beds: 51.5<br>201 – 300 beds: 28.1<br>301 – 400 beds: 25.0<br>401 – 500 beds: 33.3<br>> 500 beds: 45.3 | $\chi^2 = 11.947^{\dagger}$ |
| Hospital ICU | Present: 36.5<br>Absent: 28.6 | $\chi^2 = 0.181$ |
| MCNU | Present: 36.6<br>Absent: 34.3 | $\chi^2 = 0.066$ |
| IFU | Present: 44.4<br>Absent: 30.5 | $\chi^2 = 3.610$<br>$p = 0.0574$ |
| Experience of the attending physician | Expert: 36.7<br>High: 32.5<br>Medium: 50.0<br>Low: 26.3<br>None/Nod declared: 35.7 | $\chi^2 = 4.0867$ |

$^{\dagger}p < 0.05$ .
Source: Records of the study.
Size of the study serie: 177.

Figure 3 shows the behavior of the spontaneous closure of the fistula in each of the moments of the cohort. As noted in the preceding sections, number of hospital beds influenced upon the number of spontaneously closed fistulas observed in each moment of the cohort: at any moment, hospitals with the smallest number of beds showed the highest rates of spontaneous closures. In a complementary fashion, hospitals with the higher number of beds concentrated the higher number of patients with non-closed, active fistulas (χ^2^ = 8.481; p < 0.05; log-rank test based on the chi-square distribution).

**Figure 3.**
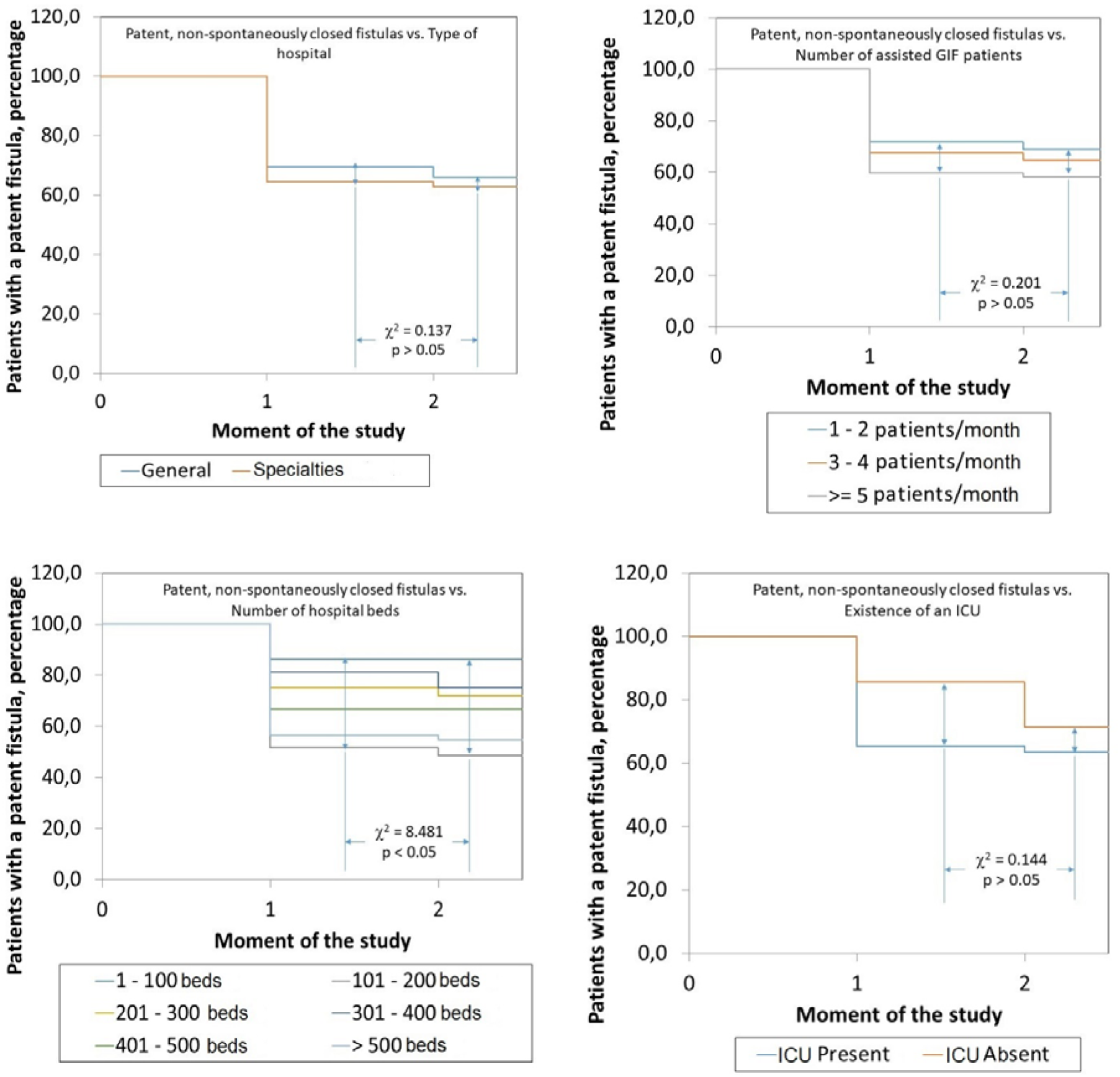
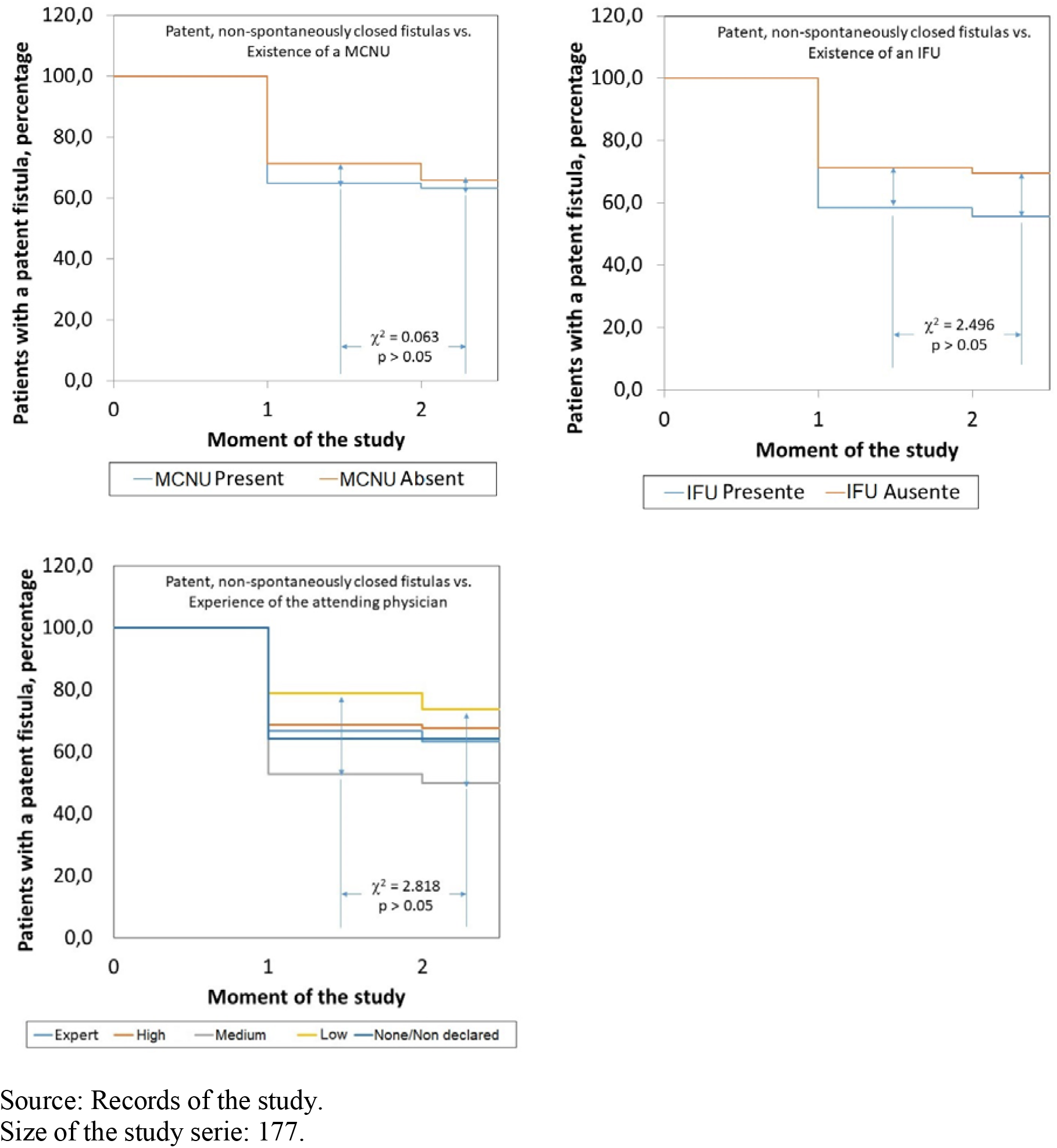
Influence of the operational characteristics of the participating hospitals upon spontaneous closure of gastrointestinal fistulas in each moment of the study. Legend: GIF: Gastrointestinal fistulas. ICU: Intensive Care Unit. MCNU: Multidisciplinary Clinical Nutrition Unit. IFU: Intestinal Failure Unit.

## DISCUSSION

The present work shows the influence of the operational characteristics of the participating hospitals upon containment, management and resolution of GIF. It is to be noticed operational characteristics explored in this essay do not exhaust all those defining the structure^‡^ and organization^§^ of a hospital, but they are expected to be those ones determining the optimal outcome of the fistula.

Three of the operational characteristics of the hospital influenced upon prognosis and evolution of GIF albeit in uneven fashion and with unequal strength: average number of patients with GIF assisted in the hospital during a month work, existence of an IFU in the center, and the number of hospital beds.

Only the number of patients with GIF assisted in the hospital in a month work influenced upon survival of the patient, and this influence was marginal (χ^2^ = 5.934; p = 0.0514). In spite of this, the association found is noticeable given the heterogeneity observed in the present study serie. Medical wisdom is built upon experience, and the more GIF cases are assisted in the institution, better actions to be performed in a given moment on given patients in order to achieve their survival (and in the process the best treatment of the fistula leading up to a favorable resolution) will be devised.^6^ However, the (favorable) association in question was not entirely linear (that is: the more the patients assisted the higher the survival rate), but inverted “U”-shaped, with the highest number of patients being discharged alive from those hospitals assisting 3 – 4 patients with FGI in a month as average regarding those assisting either 1 – 3 patients or 5 (and more) patients, thus indicating other operational characteristics of the hospital might be mediating such relationship.

On the other hand, existence of a hospital IFU also influenced marginally (χ^2^ = 3.610; p = 0.0574) upon the possibility of spontaneous closure of GIF: hospitals endowed with this organizational resource showed a trend towards a higher number of spontaneously resolved fistulas, that is, without the need of additional surgeries. An IFU made up by specialized and permanently updated personnel can be a cost-effective solution for the treatment and resolution of GIF.^7^ Similarly, as mentioned in the preceding paragraph, other operational characteristics of the hospital might be obscuring | mediating this association, and with that the influence of the IFU, thus providing an explanation for the marginal effect of this hospital characteristic.

Of all the operational characteristics considered, the number of hospital beds was the one determining both prolongation of hospital stay as well as the possibility of spontaneous closure of the fistula. The number of hospital beds could be a surrogate of the capacity of the hospital to fulfill its missions. Thus, the higher the number of beds, the better equipped | prepared the hospital will be to satisfy the goals derived from these missions. A higher number of beds would also imply the hospital is habilitated to face major surgical dramas such as those poised by GIF themselves.

In that sense, it would be anticipated a higher number of hospital beds to translate into a higher number of spontaneously closed fistulas. Such effect was observed in the present study, with hospitals with the highest number of beds achieving a higher number of fistulas spontaneously resolved. Once measures for controlling sepsis and stabilizing the inner milieu are taken, and hydroelectrolyte replenishment and nutritional support are secured, it could be then expected the fistula to spontaneously close (if there are also other factors concurring to this end). A higher number of hospital beds would then imply the availability of the resources previously mentioned in the institution for better containing, treating and solving the GIF.

Influence of the number of hospital beds upon hospital stay during containment of GIF deserves further consideration. Resolution of GIF is a time-consuming process, requiring several and concerted medical-surgical actions in order to achieve their closure.^8^ It is then immediate resolution of GIF will consume a significant quota of admission time in the hospital for conducting (and assessing the impact) of the previously mentioned actions. If the number of hospital beds behaves as a surrogate of the sanitary and technological complexity of the hospital, then the higher number of beds would imply (by force) more prolonged hospital stays. That was the case in the present study, where a positive and significant association was found between the number of hospital beds and prolongation of hospital stay.

As described in other parts of this text, the relationship between the number of hospital beds and prolonged hospital stay was “U”-shaped instead of linear, and this would point to another factors of different type (structural | organizative | cultural) modelling such association. A better understanding of the influence of the operational characteristics of the hospital upon containment and resolution of GIF will be obtained after examining the surgical practices medical care teams adopt in GIF patients assisted in the surveyed centers. Regarding this point, two tentative hypotheses can be put forward: actions are taken for closing the fistulas in “small” hospitals that are not duly prepared for it, unnecessarily prolonging the hospital stay without achieving a resolution neither definitive nor cost-effective. On the other hand, it is possible that in “medium” hospitals in which “shortened” hospital stays are seen medical care teams are dealing with intestinal leakages instead of established fistulas, leakages that would evolve towards the spontaneous closure in a few days with a minimal (or for the same reason reduced) quota of actions and resources.

## CONCLUSIONS

Treatment and resolution of GIF might be dependent upon some of the operational characteristics of the hospital, among them, the number of hospital beds, the average number of patients with GIF assisted in a month work, and the existence of a hospital IFU. However, such associations would be mediated, on one hand, by the heterogeneity of the study serie, and the existence of another structural | organizative | cultural factors related with the way medical care teams face GIF locally.

## Supporting information

Statement from the San Javier Hospital IRB

## Data Availability

Interested researchers and parties are invited to contact the authors for guidelines pertaining access, use and reuse of the data collected during the study.

## Further extensions

Influence of surgical practices medical care teams adopt within the hospital to contain, treat and successfully solve GIF will be examined in succeeding essays thus extending the results presented in this report.

## Footnotes

* Available at: http://www.redcap.org.

† A photocopy of the ruling by the Ethics Committee of the San Javier Hospital is included as a supplemental material.

‡ Structure: Sum of the sanitary facilities, installed equipment, assigned budgets, and hired human resources.

§ Organization: Content and purposes of the procedures being conducted within the institution. Organization = Structure + Procedures.

## Notes

### Competing Interest Statement

The authors have declared no competing interest.

### Funding Statement

The authors of the "Fistula Day" Project did not apply for grants or by the same token financial support for its conduction.

### Author Declarations

The present essay is a follow-up to a previous publication with the design and an exploratory analysis of the results of the "Fistula Day". Accordingly, the protocol followed by local surveyors during the "Fistula Day" was drafted according with "Good Clinical Practices". Identity and rights of the surveyed patients were also protected. The patients (and by extension their caretakers) were informed about the purposes of the research, and the non-invasive nature of the procedures. Collected data were adequately preserved in order to ensure anonymity and confidentiality. Aggregated data were used in the interpretation of the results and the realization of statistical inferences. Informed consent was obtained from the patient before inclusion in the cohort. Local conduction of the activities foreseen in the "Fistula Day" was authorized and supervised by the hospital Committees of Ethics after presentation, review and approval of the research protocols. The researchers entrusted with the conduction of the "Fistula Day" Project presented the protocol "Current status of the postoperative fistula of digestive tract; multicentric, multinational study. DAY OF THE FISTULA" before the Ethics Committee of the San Javier Hospital (city of Guadalajara, State of Jalisco, Mexico) for review and approval. A ruling was emitted on April 11th, 2018 by Dr. Eduardo Razon Gutierrez, acting Director of the Ethics Committee, with the approval of the research protocol and the authorization for the conduction of the "Fistula Day" Project.

